# Predicting clinical outcomes in the Machine Learning era: The Piacenza score a purely data driven approach for mortality prediction in COVID-19 Pneumonia

**DOI:** 10.1101/2021.03.16.21253752

**Authors:** Geza Halasz, Michela Sperti, Matteo Villani, Umberto Michelucci, Piergiuseppe Agostoni, Andrea Biagi, Luca Rossi, Andrea Botti, Chiara Mari, Marco Maccarini, Filippo Pura, Loris Roveda, Alessia Nardecchia, Emanuele Mottola, Massimo Nolli, Elisabetta Salvioni, Massimo Mapelli, Marco Agostino Deriu, Dario Piga, Massimo Piepoli

## Abstract

**Background:** Several models have been developed to predict mortality in patients with COVID-19 pneumonia, but only few have demonstrated enough discriminatory capacity. Machine-learning(ML) algorithms represent a novel approach for data-driven prediction of clinical outcomes with advantages over statistical modelling. We developed the Piacenza score, a ML-based score, to predict 30-day mortality in patients with COVID-19 pneumonia.

**Methods:** 852 patients (mean age 70years, 70%males) were enrolled from February to November 2020. The dataset was randomly splitted into derivation and test. The Piacenza score was obtained through the Naïve Bayes classifier and externally validated on 86 patients. Using a forward-search algorithm the following six features were identified: age; mean corpuscular haemoglobin concentration; PaO^2^ /FiO^2^ ratio; temperature; previous stroke; gender. In case one or more of the features are not available for a patient, the model can be re-trained using only the provided features.

We also compared the Piacenza score with the 4C score and with a Naïve Bayes algorithm with 14 variables chosen a-priori.

**Results:** The Piacenza score showed an AUC of 0.78(95% CI 0.74-0.84, Brier-score 0.19) in the internal validation cohort and 0.79(95% CI 0.68-0.89, Brier-score 0.16) in the external validation cohort showing a comparable accuracy respect to the 4C score and to the Naïve Bayes model with a-priori chosen features, which achieved an AUC of 0.78(95% CI 0.73-0.83, Brier-score 0.26) and 0.80(95% CI 0.75-0.86, Brier-score 0.17) respectively.

**Conclusion:** A personalized ML-based score with a purely data driven features selection is feasible and effective to predict mortality in patients with COVID-19 pneumonia.

## Introduction

Despite measureless efforts to limit COVID-19 pandemic spread, over 100 million people have been confirmed positive for SARS-CoV-2 and more than 2 million people have died from the virus worldwide, as of 10 February 2021.^1^

While these numbers rapidly increase day by day, hospitals have been receiving requests beyond capacity and face extreme challenges concerning a sharp increase in the demand for medical resources and the shortage of hospital beds and critical care equipment for the timely treatment of ill patients.

Additionally, the clinical spectrum of SARS-CoV-2 infections ranges from asymptomatic status to severe viral pneumonia with respiratory failure and even death, making reliable and successful patient triaging challenging.^2^

Data from epidemiological studies suggests that severe illness occurs in approximately 20% of the patients and that older age, coexisting medical conditions and cardiovascular risk factors are associated with worse prognosis^3-4^.

In this scenario, identification of key patients’ variables driving COVID-19 prognosis is of paramount importance to assist physicians in early predicting pathology trajectory, and improving patient outcomes. To date several prognostic models, combining clinical and laboratory parameters, have been proposed but they included mainly patients from the first wave of COVID-19 infection. This may cause a risk of bias, making these models unsuitable for clinical decision in daily practice^5-6^.

The increasing use of electronic health-care record (EHR) systems has increased the availability of a large amount of data suitable for Machine Learning (ML) analysis. The latter has already proven its potential to support clinical decisions in many medical fields, including COVID-19 pandemics.^7-8^

Therefore, the aim of the present study was to develop and validate a new score (named Piacenza score) to predict the prognosis of COVID-19 pneumonia, based on a ML technique with a purely data-driven selection of prognostic features collected at hospital admission.

We hypothesized that a ML score based on data-driven selection, differently from inference statistics, could capture non-linear relationships among clinical features without human-biased intervention and could predict mortality for individual patients more accurately than the currently available risk scores.

The Piacenza score was specifically designed to be an easy, fast, versatile, fair, open, and user-friendly tool. To reach this goal, a web-based calculator of the score has been released, available at (https://covid.7hc.tech.).

This calculator can be used by clinicians to estimate an individual hospitalized patient’s risk of 30-day mortality. Moreover, our score is intended to be as much generalizable as possible but at the same time customizable to the single patient and flexible to be applied with different variables.

Thus, if one or more features needed to compute the Piacenza score are not available in a single patient, the clinician anyway can receive a risk stratification score by providing the value of different variables from a subset proposed by the system. In this case, the ML model is retrained to compute a mortality risk tailored to the patient leading to a highly customizable score.

## Methods

### Population and collected data

The study was conducted at Guglielmo Saliceto Hospital which serves a population of about 300’000 people in the area of Piacenza, Emilia Romagna (North Italy), that represents the second region in Italy for number of COVID-19 deceased persons (6219 at the date of December 7th, 2020).

The present research retrospectively analysed the electronic health records (EHR) of a cohort of 852 patients, diagnosed with COVID-19 pneumonia according to the WHO interim guidance, admitted to the hospital from February to November 2020.

COVID-19 infection was diagnosed by a positive result on a reverse-transcriptase–polymerase-chain-reaction (RT-PCR) assay of a specimen collected on a nasopharyngeal swab. Pregnant women, children (<18 years) and patients with negative RT-PCR assay were excluded from the study as well as patients presenting with shock and coma.

Data collected in EHR included patients’ demographic information, comorbidities, triage vitals, laboratory tests and outcomes (including length of stay, discharge, readmission, and mortality). Routine blood examinations at admission comprised complete blood count, coagulation profile, serum biochemical tests (including renal and liver function, creatine kinase, lactate dehydrogenase, electrolytes, C-reactive protein). A total of 62 patient’s characteristics were considered in the score design and development. The study protocol was approved by the local committee on human research.

### Criteria for discharge and outcome

The criteria for discharge were at the discretion of the caregiver physician. Mostly, criteria encompassed absence of fever for at least three days, substantial clinical improvement including clinical remission of symptoms and two throat-swab samples negative for SARS-CoV-2 RNA obtained at least 24h apart. The primary outcome was 30-day in-hospital mortality.

### Piacenza score design

The Piacenza score is a ML-based COVID-19 mortality risk predictor. It was implemented using a Naïve Bayes approach, which is a probabilistic classifier describing the dependence from the outcome of each variable characterizing the patient, taken separately from the others. The Naïve Bayes algorithm was chosen due to the following advantages: (i)it provides a probability of the final outcome, which thus represents the mortality risk; (ii)it can handle both categorical and continuous features; (iii)it can handle missing values, thus providing a mortality risk even when not all features of a patient are available. Moreover, it proved a successful approach in predicting clinical outcomes in several medical scenarios^9-10^.

### Derivation and test cohorts

The available EHR of 852 patients was randomly split in derivation (70%) and test (30%) cohorts. The derivation cohort was first used to select, among the considered 62 patient’s features, the most significant ones and then to train the Naïve Bayes classifier considering only the best top predictors, while the predictive ability of the estimated model was assessed on the test cohort.

### Piacenza score development, optimization and identification of variable importance

The Piacenza score has been developed and tailored to: (i) minimize the number of clinical variables to be ingested and (ii) to maximize the overall prediction performance (i.e., in terms of maximization of the area under the receiver operating characteristic curve (AUC)) and patient stratification ability. The most significant patient’s features were identified through the so called forward-search approach^11^.

The forward search approach is a purely data-driven dimensionality reduction technique able to identify, given a large set of input features, the minimum combination of those features which maximize the performance metrics associated to the machine learning algorithm. The forward search approach was here employed to reduce the number of patient variables from 62 to the six most relevant used to train the Naïve Bayes classifier.

### Piacenza score evaluation and metrics

The test cohort was used to assess the performance of the Piacenza score. In order to increase the statistical significance of the results, bootstrapping was used to randomly generate 100 test sets from the original test cohort. Moreover, an external validation cohort has been considered to further validate the Piacenza score performance. The external validation cohort consisted of data from 86 COVID-19 patients enrolled at Centro Cardiologico Monzino Hospital (Milan, Italy).

The Piacenza score performances were evaluated in terms of discrimination and calibration capabilities. The discrimination ability was determined by computing the receiver operating characteristic (ROC) curve on the test cohort and the associated AUC, together with its 95% confidence interval (CI).

The calibration ability was derived by the so-called calibration plots, which compare observed and predicted outcomes with associated uncertainties. The Brier index was used to evaluate the ability of ML to stratify and predict observed outcomes. The Brier index is defined as the mean-squared difference between the observed and predicted outcomes and ranges from 0 to 1, with 0 representing the best calibration. Finally, the variable relative importance has been quantified for the identified 6 most relevant patient features. The relative importance is a comparative measure of the patient’s feature’s weight in determining the Piacenza risk score.

### Usability, flexibility and customization

Additional steps were performed to make the Piacenza score flexible and customizable.

A user-friendly web site was designed and developed to enable a fast and easy use of the tool by the final user (i.e., the physician).

Regarding the customization properties to the Piacenza score, we added a personalized version of the algorithm inside the website, which enables an optimized computation of the mortality risk score for a single patient, when some variables used by the Piacenza score are not available. In this case, the Naïve Bayes classifier is re-trained over the same derivation cohort but using a different set of patient’s characteristics. More specifically, the following 14 variables have been chosen a priori by the physician because their association with mortality in COVID-19 pneumonia: age; gender; diabetes; length of symptoms before hospital admission; systolic blood pressure; respiratory rate (RR); PaO2/FiO2 (P/F) ratio; platelets and eosinophils count; neutrophils to lymphocytes ratio (NLR); CRP; direct bilirubin; creatinine; lactate dehydrogenase (LDH). Finally, we compared the performance of the Piacenza Score with the above mentioned “clinical” Naïve Bayes classifier.

### Comparison with conventional risk models

To further assess the performance of the Piacenza score, we compared it with the 4C mortality score, which considers the following predictors: age; gender; number of comorbidities; respiratory rate (RR); peripheral oxygen saturation (SO2); level of consciousness (Glasgow coma scale); urea level; C reactive protein (CRP). The same test cohort used to test the Piacenza score was employed.

### Statistical analysis

Categorical variables are reported as count (%) and continuous variables as mean (standard deviation, SD). A two-sided p-value (p) < 0.05 was considered statistically significant. We used Fisher’s exact test to assess differences between binary variables and Welch’s 2-sample t-test to assess differences between continuous variables. The overall implementation of all codes for the machine learning score and analysis tools was performed in Python 3.7.4 environment.

## Results

### Patient characteristics and events

852 patients with SARS-CoV-2 pneumonia were hospitalized during the study period, among which 242 (28%) were admitted to the intensive care unit (ICU). The mean age of the patients was 70 (±14) and 599 (70%) were male. Comorbidities were present in 602 patients (71%): mainly arterial hypertension (59%), dyslipidaemia (24%) and diabetes (18%). The mean time between onset of symptoms and hospital admission was 6.5 days (±3.9). Fever (94.5%), dyspnea (63.7%) and cough (46.8%) were the most common symptoms on admission. 293 patients (34%) died within 30 days after hospital admission, the median time from hospital admission to discharge or death was 9 days. The comparison of clinical characteristics between survivors and non-survivors showed that the latter were older (p<0.001); had a higher prevalence of Hypertension and cerebrovascular disease (p<0.001); longer symptom duration(p<0.001); higher respiratory rate(p<0.001); lower Sp02(p<0.001); Pao2/FiO2 ratio(p<0.001); and systolic blood pressure on admission (p=0.019).

Major laboratory markers were tracked on admission. Specifically, lactate dehydrogenase (LDH), creatine kinase (CK), cholinesterase (CH), creatinine, and glycemia were significantly higher in non-survivors than survivors (p<0.001). Non survivors had significantly lower lymphocytes and eosinophils percentage and Red Blood cells count as well as HB, MCHC and HCT values(p<0.001). Furthermore, non survivors showed significantly higher levels of inflammatory biomarkers such as neutrophils count, CPR and NLR values (p<0.001). Other differences in laboratory findings among the two groups are summarized in Table 1.

**TABLE 1:**
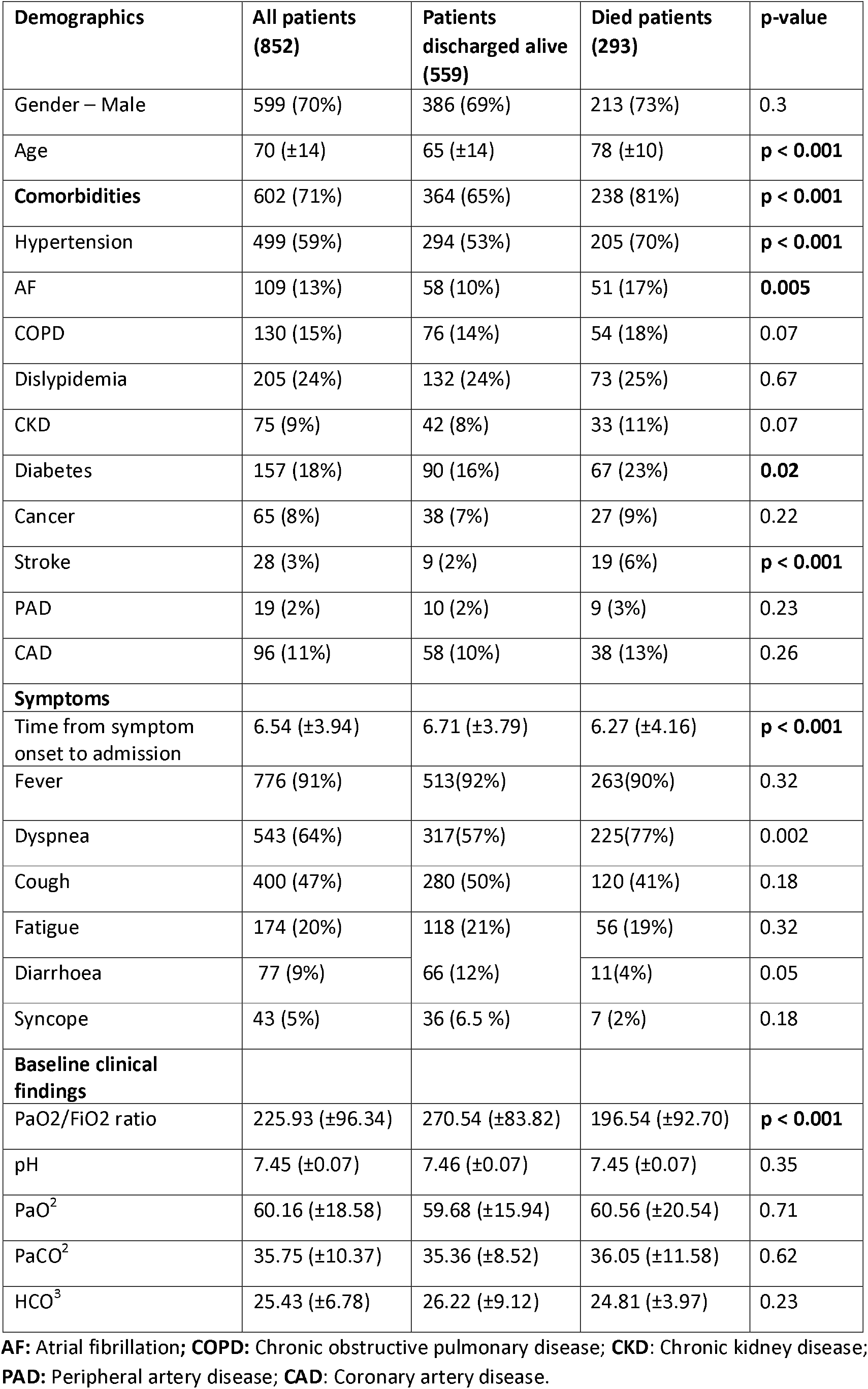
Study population characteristics. Data are n; mean (%) or standard deviation(sd). Columns refers to overall study sample and compare the groups of survivors and non-survivors. p-value refers to either Student’s t-test or χ2 test.

| Demographics | All patients (852) | Patients discharged alive (559) | Died patients (293) | p-value |
| --- | --- | --- | --- | --- |
| Gender – Male | 599 (70%) | 386 (69%) | 213 (73%) | 0.3 |
| Age | 70 ($\pm 14$) | 65 ($\pm 14$) | 78 ($\pm 10$) | **p < 0.001** |
| **Comorbidities** | 602 (71%) | 364 (65%) | 238 (81%) | **p < 0.001** |
| Hypertension | 499 (59%) | 294 (53%) | 205 (70%) | **p < 0.001** |
| AF | 109 (13%) | 58 (10%) | 51 (17%) | **0.005** |
| COPD | 130 (15%) | 76 (14%) | 54 (18%) | 0.07 |
| Dislipidemia | 205 (24%) | 132 (24%) | 73 (25%) | 0.67 |
| CKD | 75 (9%) | 42 (8%) | 33 (11%) | 0.07 |
| Diabetes | 157 (18%) | 90 (16%) | 67 (23%) | **0.02** |
| Cancer | 65 (8%) | 38 (7%) | 27 (9%) | 0.22 |
| Stroke | 28 (3%) | 9 (2%) | 19 (6%) | **p < 0.001** |
| PAD | 19 (2%) | 10 (2%) | 9 (3%) | 0.23 |
| CAD | 96 (11%) | 58 (10%) | 38 (13%) | 0.26 |
| **Symptoms** |  |  |  |  |
| Time from symptom onset to admission | 6.54 ($\pm 3.94$) | 6.71 ($\pm 3.79$) | 6.27 ($\pm 4.16$) | **p < 0.001** |
| Fever | 776 (91%) | 513(92%) | 263(90%) | 0.32 |
| Dyspnea | 543 (64%) | 317(57%) | 225(77%) | 0.002 |
| Cough | 400 (47%) | 280 (50%) | 120 (41%) | 0.18 |
| Fatigue | 174 (20%) | 118 (21%) | 56 (19%) | 0.32 |
| Diarrhoea | 77 (9%) | 66 (12%) | 11(4%) | 0.05 |
| Syncope | 43 (5%) | 36 (6.5 %) | 7 (2%) | 0.18 |
| **Baseline clinical findings** |  |  |  |  |
| PaO<sub>2</sub>/FiO<sub>2</sub> ratio | 225.93 ($\pm 96.34$) | 270.54 ($\pm 83.82$) | 196.54 ($\pm 92.70$) | **p < 0.001** |
| pH | 7.45 ($\pm 0.07$) | 7.46 ($\pm 0.07$) | 7.45 ($\pm 0.07$) | 0.35 |
| PaO<sub>2</sub> | 60.16 ($\pm 18.58$) | 59.68 ($\pm 15.94$) | 60.56 ($\pm 20.54$) | 0.71 |
| PaCO<sub>2</sub> | 35.75 ($\pm 10.37$) | 35.36 ($\pm 8.52$) | 36.05 ($\pm 11.58$) | 0.62 |
| HCO<sub>3</sub> | 25.43 ($\pm 6.78$) | 26.22 ($\pm 9.12$) | 24.81 ($\pm 3.97$) | 0.23 |

**AF:** Atrial fibrillation; **COPD:** Chronic obstructive pulmonary disease; **CKD:** Chronic kidney disease; **PAD:** Peripheral artery disease; **CAD:** Coronary artery disease.

**TABLE 2:** Laboratory findings on admission Data are n; mean and standard deviation. Columns refers to overall study sample and compare the groups of survivors and non-survivors. P value refers to either Student’s t-test or χ2 test.

| Laboratory parameters | All patients (852) | Patients discharged alive (559) | Died patients (293) | p-value |
| --- | --- | --- | --- | --- |
| GLUCOSE mg/dl | 145 ($\pm 66$) | 137 ($\pm 59$) | 159 ($\pm 76$) | **p < 0.001** |
| UREA mg/dl | 57 ($\pm 40$) | 47 ($\pm 24$) | 76 ($\pm 54$) | **p < 0.001** |
| CREATININE mg/dl | 1.24 ($\pm 0.90$) | 1.06 ($\pm 0.54$) | 1.59 ($\pm 1.27$) | **p < 0.001** |
| SODIUM mEq/l | 137 ($\pm 8$) | 137 ($\pm 8$) | 137 ($\pm 7$) | 0.24 |
| POTASSIUM mEq/l | 4.17 ($\pm 0.55$) | 4.14 ($\pm 0.49$) | 4.24 ($\pm 0.65$) | **0.04** |
| CHLORIDE mEq/l | 99.26 ($\pm 7.21$) | 98.84 ($\pm 7.19$) | 100.05 ($\pm 7.17$) | **0.02** |
| TOTAL BILIRUBIN mg/dl | 0.75 ($\pm 0.48$) | 0.72 ($\pm 0.35$) | 0.82 ($\pm 0.66$) | **0.02** |
| DIRECT BILIRUBIN mg/dl | 0.22 ($\pm 0.60$) | 0.21 ($\pm 0.69$) | 0.25 ($\pm 0.37$) | 0.31 |
| AST U/L | 61 ($\pm 84$) | 53 ($\pm 37$) | 79 ($\pm 136$) | **0.004** |
| ALT U/L | 48 ($\pm 70$) | 47 ($\pm 44$) | 48 ($\pm 103$) | 0.90 |
| LDH U/L | 430 ($\pm 220$) | 391 ($\pm 160$) | 509 ($\pm 292$) | **p < 0.001** |
| CK U/L | 300 ($\pm 637$) | 231 ($\pm 387$) | 429 ($\pm 932$) | **p < 0.001** |
| AMILASE U/L | 73 ($\pm 48$) | 69 ($\pm 37$) | 80 ($\pm 63$) | **0.013** |
| LIPASE U/L | 47 ($\pm 72$) | 43 ($\pm 46$) | 56 ($\pm 105$) | 0.06 |
| SERUM CHOLINESTERASE U/L | 6275 ($\pm 1858$) | 6674 ($\pm 1763$) | 5576 ($\pm 1812$) | **p < 0.001** |
| WBC $\times 10^3/\mu\text{l}$ | 8.12 ($\pm 4.68$) | 7.86 ($\pm 4.72$) | 8.63 ($\pm 4.56$) | **0.02** |
| RBC $\times 10^6/\mu\text{l}$ | 4.69 ($\pm 0.72$) | 4.79 ($\pm 0.68$) | 4.51 ($\pm 0.77$) | **p < 0.001** |
| HB gr/dl | 13.59 ($\pm 1.91$) | 13.83 ($\pm 1.72$) | 13.14 ($\pm 2.16$) | **p < 0.001** |
| HCT % | 41.84 ($\pm 5.70$) | 42.37 ($\pm 5.34$) | 40.83 ($\pm 6.22$) | **p < 0.001** |
| MCV fl | 89.74 (±6.66) | 89.18 (±5.62) | 90.80 (±8.19) | **0.003** |
| MCH pg | 29.13 (±2.38) | 29.05 (±2.12) | 29.28 (±2.80) | 0.23 |
| MCHC gr/dl | 32.43 (±1.36) | 32.56 (±1.15) | 32.17 (±1.66) | **p < 0.001** |
| PLATELETS x10<sup>3</sup>/μl | 217.75 (±117.90) | 221.08 (±127.10) | 211.41 (±97.72) | 0.22 |
| RDW % | 13.65 (±1.65) | 13.27 (±.27) | 14.29 (±1.99) | **p < 0.001** |
| NEUTROPHILS % | 77.45 (±11.57) | 75.81 (±11.75) | 80.56 (±10.55) | **p < 0.001** |
| LYMPHOCYTES % | 15.17 (±9.20) | 16.48 (±9.45) | 12.67 (±8.15) | **p < 0.001** |
| MONOCYTES % | 6.89 (±4.30) | 7.16 (±4.01) | 6.36 (±4.76) | **0.015** |
| EOSINOPHILS % | 0.32 (±0.91) | 0.38 (±1.05) | 0.20 (±0.54) | **0.001** |
| LYMPHOCYTES x10<sup>3</sup>/μl | 1.09 (±0.99) | 1.15 (±0.94) | 0.98 (±1.09) | **0.029** |
| MONOCYTES x10<sup>3</sup>/μl | 0.51 (±0.41) | 0.52 (±0.35) | 0.51 (±0.51) | 0.77 |
| EOSINOPHILS x10<sup>3</sup>/μl | 0.02 (±0.07) | 0.03 (±0.08) | 0.02 (±0.05) | **0.04** |
| NEUTROPHILS x10<sup>3</sup>/μl | 6.41 (±3.72) | 6.05 (±3.41) | 7.11 (±4.15) | **p < 0.001** |
| PT sec | 15.84 (±8.38) | 15.07 (±5.83) | 17.03 (±11.11) | **0.02** |
| PROTHROMBIN ACTIVITY % | 68.40 (±15.96) | 69.86 (±14.38) | 66.27 (±17.82) | **0.009** |
| INR | 1.40 (±0.76) | 1.34 (±0.65) | 1.51 (±0.93) | **0.01** |
| PTT sec | 31.70 (±5.74) | 31.32 (±4.48) | 32.29 (±7.22) | 0.08 |
| PTT -Ratio | 1.02 (±0.19) | 1.00 (±0.14) | 1.04 (±0.25) | 0.06 |
| CPR mg/dl | 11.19 (±8.55) | 9.85 (±7.88) | 13.74 (±9.17) | **p < 0.001** |
| NLR | 7.99 (±6.74) | 6.78 (±5.04) | 10.27 (±8.68) | **p < 0.001** |

**AST:** Aspartate aminotransferase; **ALT:** Alanine aminotransferase; **LDH:** Lactate Dehydrogenase; **CK:** Creatine kinase **WBC:** White blood cell counts; **RBC:** Red blood cell count; **HB:** haemoglobin; **HCT:** hematocrit; **MCV:** Mean Corpuscular Volume; **MCH:** Mean Corpuscular Hemoglobin; **MCHC:** Mean Corpuscular Hemoglobin Concentration; **RDW:** Red Cell Distribution Width; **PT:** prothrombin time; **PTT:** Partial Thromboplastin Time; **CRP:** C-reactive protein; **NLR:** neutrophil lymphocyte ratio.

### Significant predictors and Piacenza score

Using the forward-search algorithm, the following six most important predictors at hospital admission are identified and used to compute the Piacenza score: age; mean corpuscular haemoglobin concentration (MCHC); P/F ratio; temperature; previous cerebrovascular stroke; gender.

The median of the ROC curve over 100 test cohorts (generated through bootstrapping) is reported in Figure 1. The corresponding median of the AUC is equal to 0.78 (95% CI 0.74-0.84).

**Figure 1:**
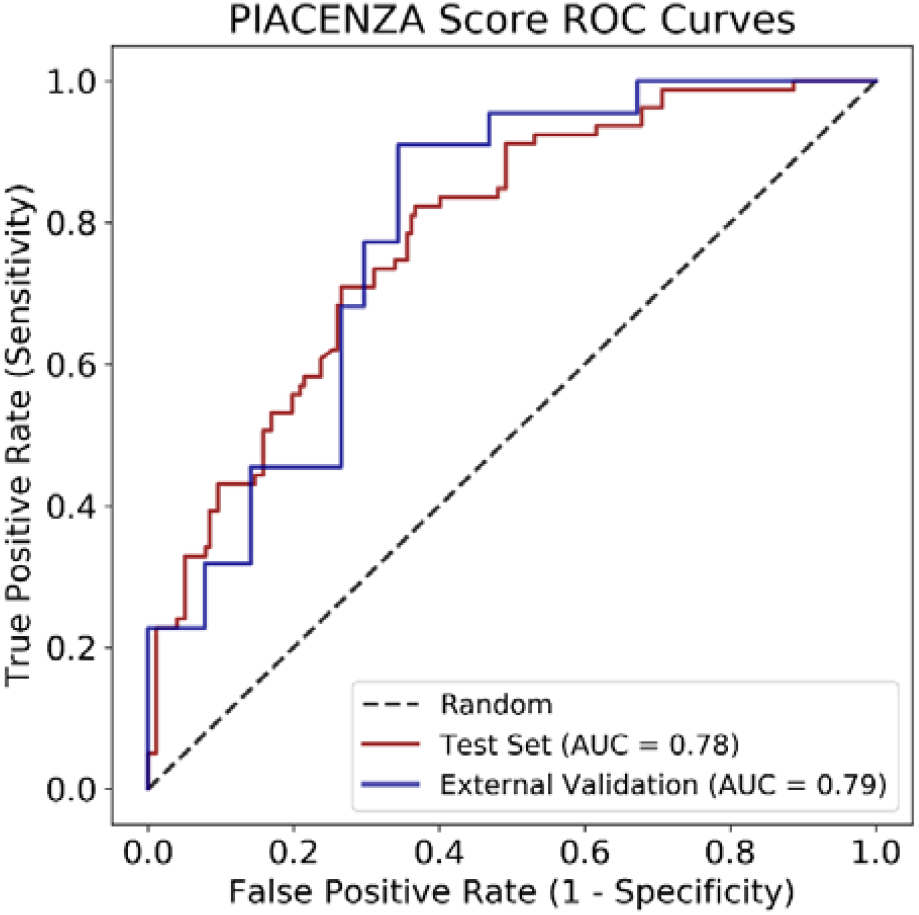
ROC curves obtained by evaluating the Piacenza score (red curve) on the test cohort and the Piacenza score on the external validation cohort of 86 patients from a different hospital (blue curve). The ROC curves shown in the figure are the median over 100 bootstrapped repetitions of the two test sets.

**Figure 2:**
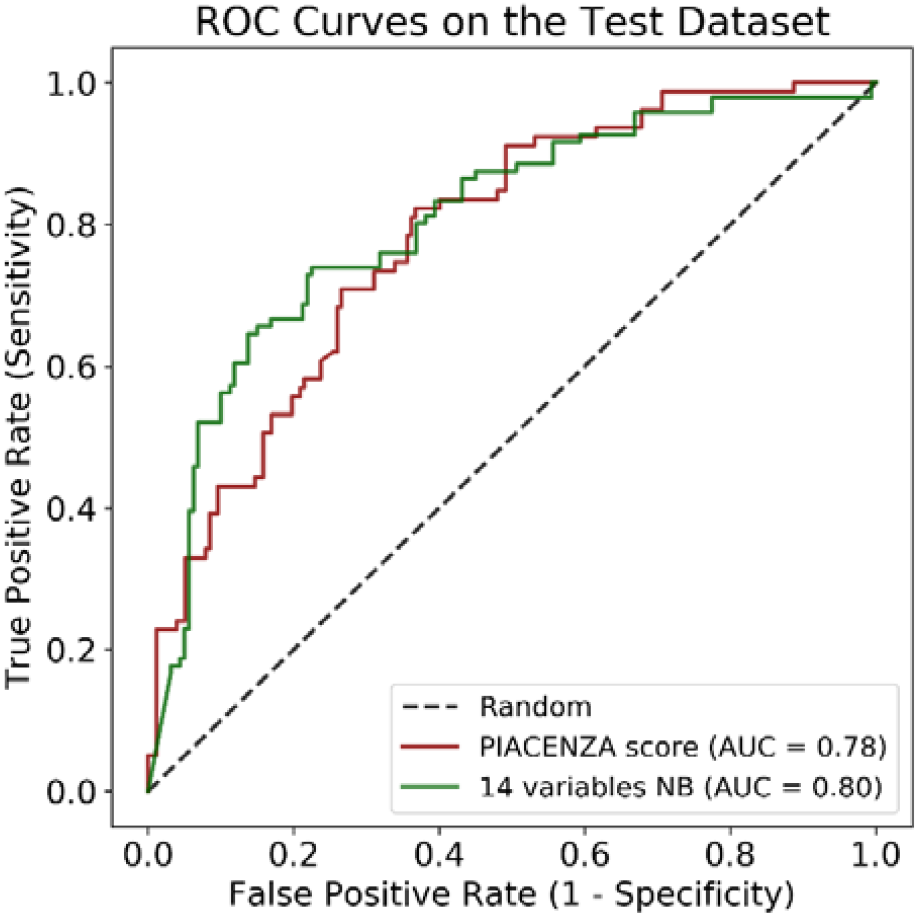
ROC curves obtained by evaluating the Piacenza score (red curve) and the Naïve Bayes model trained with 14 manually-chosen features (green curve). The ROC curves are a median over the 100 bootstrapped repetitions of the same test set.

The calibration plot of Piacenza score over the range of risk showed a Brier Score of 0.19. The risk deciles are grouped into three levels: low risk (first to fifth deciles); intermediate risk (sixth to eighth deciles) and high risk (ninth and tenth deciles). A gradual and progressive increase in absolute event rates was observed across risk classes for all the Piacenza scores (death: 14% [18/125] in low-risk deciles vs 36% [27/75] in intermediate-risk deciles vs 66% [33/50] in high-risk deciles).

From the computed calibration plot, we can observe that the mortality risk is underestimated only in the first few deciles, while in the higher deciles the risk is slightly overestimated (Figure 3A-D).

**Figure 3:**
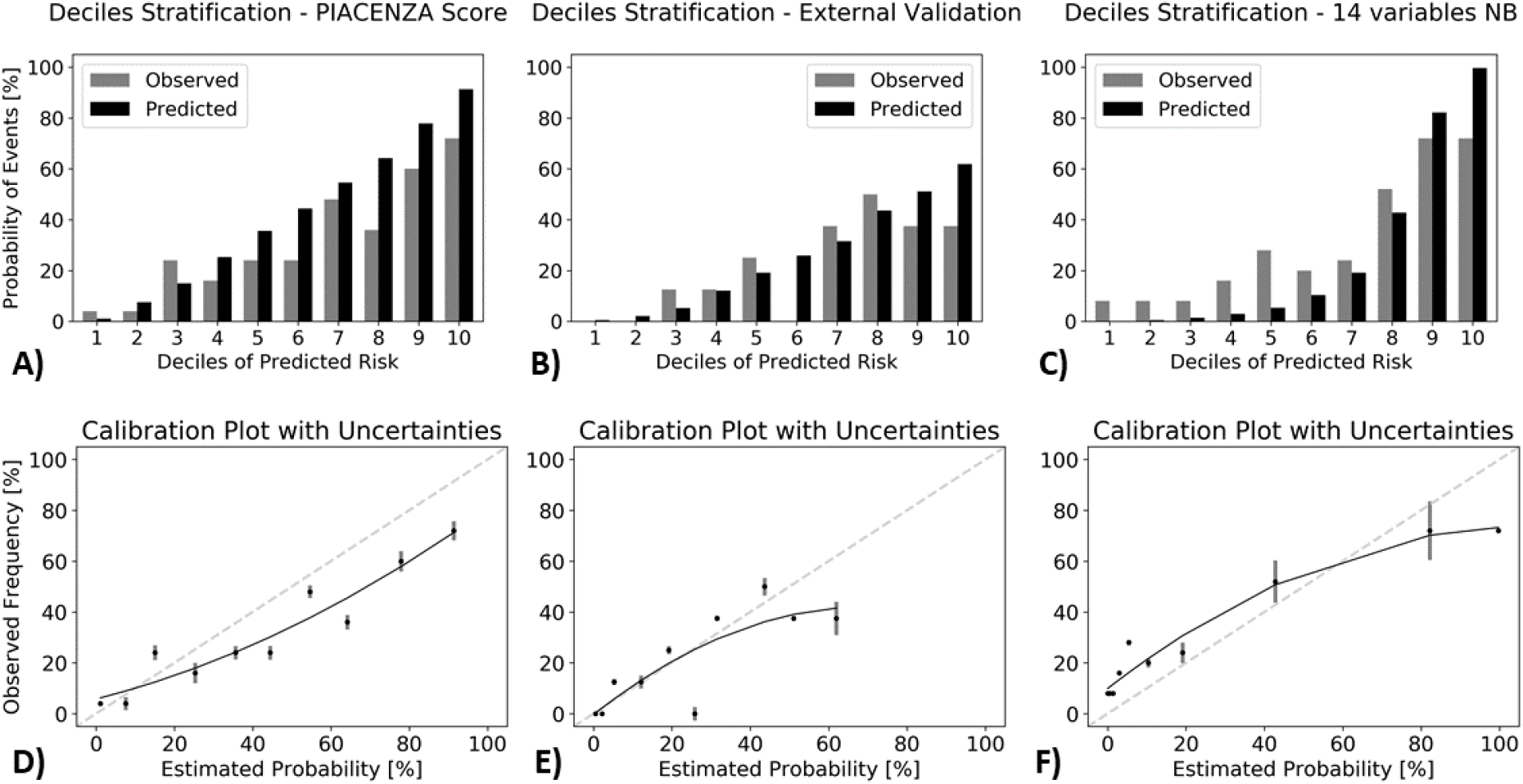
Risk of observed death according to deciles of event probability based on the Piacenza score (A), the Piacenza score on the external validation dataset (B) and the Naïve Bayes model trained with 14 manually-chosen features (C). For each single case the corresponding calibration plots with standard deviations calculated over the deciles are also shown below each respective figure (D, E, F).

Regarding the relative importance of each features taken independently from the others Age were the most important features to predict death followed by MCHC, P/F ratio, previous cerebrovascular stroke, gender and temperature (Figure 4).

**Figure 4:**
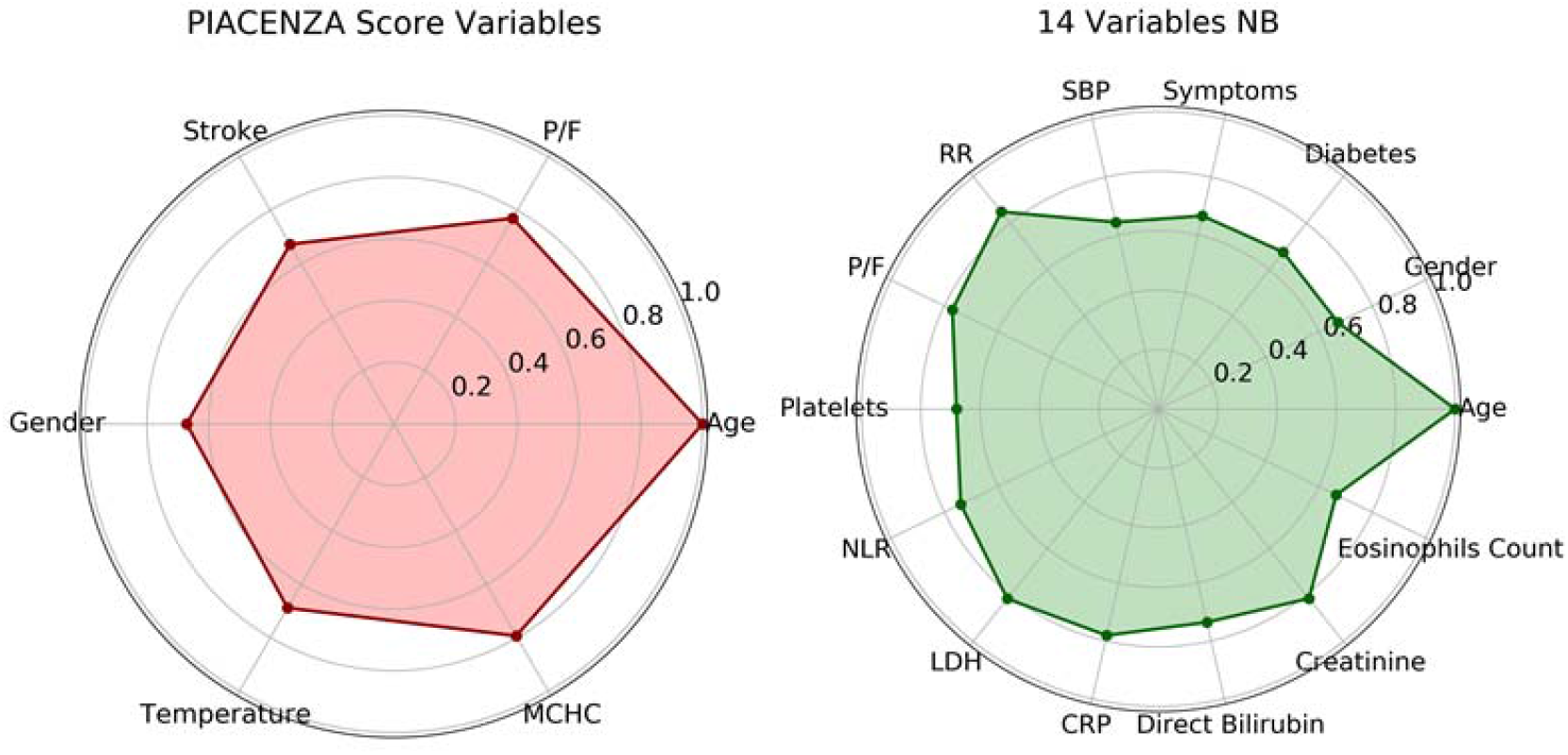
Radar plot for the 6 Piacenza score predictors of death and for the 14 manually-chosen features, showing their relative importance. (MCHC = mean corpuscular haemoglobin concentration, CRP = c-reactive protein, LDH = lactate dehydrogenase, NLR = neutrophils to lymphocytes ratio, RR = respiratory rate, SBP = systolic blood pressure). All feature importance is scaled with respect to the most important one.

### External validation

The corresponding median of the AUC in external validation cohort was 0.79 (95% CI 0.68-0.89) with a Brier score of 0.16(Figure 1).

The calibration plot is reported in Figure 3B and showed again a gradual and progressive increase in absolute event rates across risk classes (death 10% [4/40] in low-risk deciles vs 29% [7/24] in intermediate-risk deciles vs 38% [6/16] in high-risk deciles).

### Comparison with 4C mortality score and Naïve Bayes model using manually-chosen features

The median of the AUC was 0.78 (95% CI; 0.73-0.83) for the 4C score when evaluated on the test cohort the corresponding Brier score was equal to 0.26 (Figure 5).

**Figure 5:**
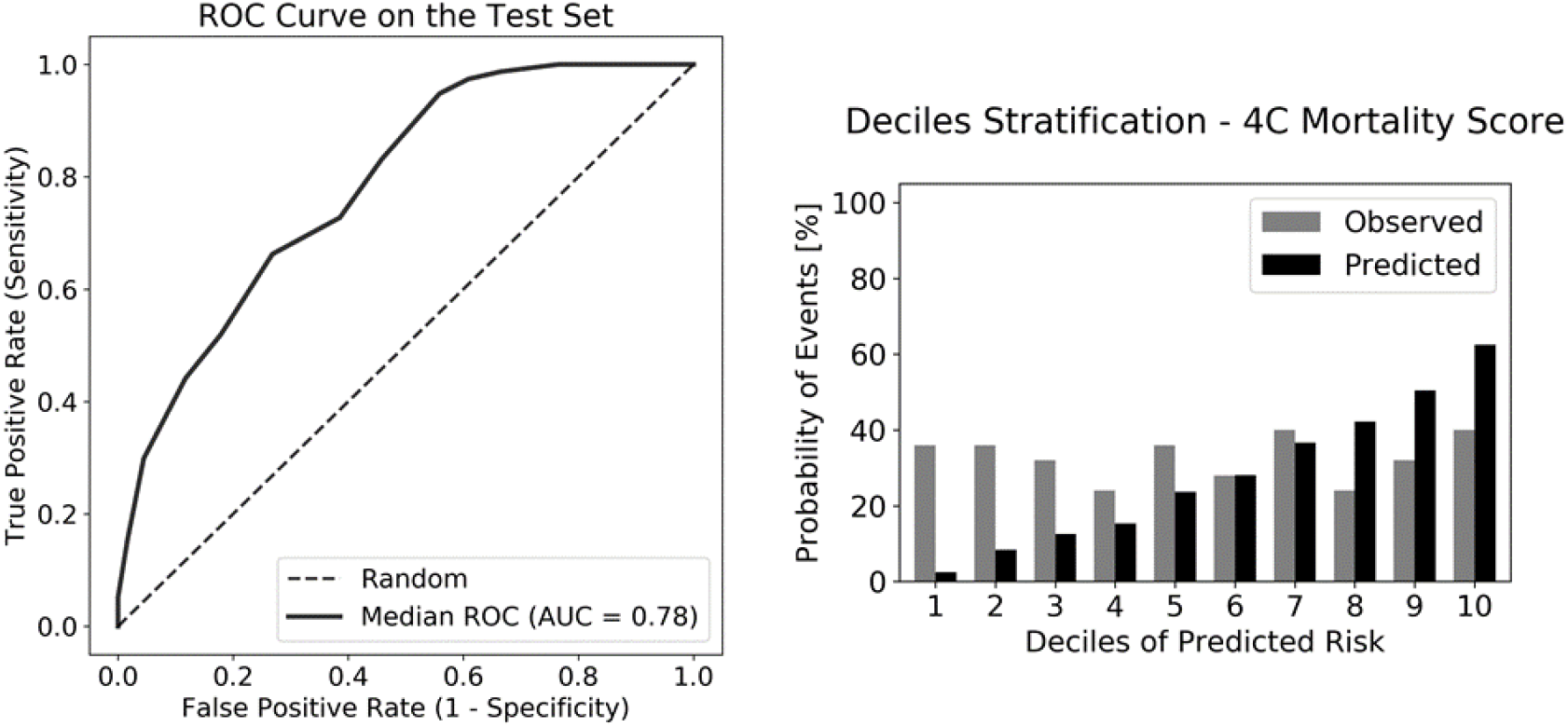
Performances of 4C mortality score (both in terms of discrimination and calibration abilities) calculated on the test cohort.

The Naïve Bayes model with 14 features chosen manually based on clinicians’ experience achieved an AUC of 0.80 (95% CI 0.75-0.86) with a Brier score of 0.17(Figure 2).

The observed mortality increased gradually and progressively for the Naïve Bayes model with manually-chosen features: death 14% [17/125] in low-risk deciles vs 32% [14/75] in intermediate-risk deciles vs 72% [36/50] in high-risk deciles, but not for 4C score: death 33% [41/125] in low-risk deciles vs 31% [23/75] in intermediate-risk deciles vs 36% [18/50] high-risk deciles. Both scores achieved a satisfactory patients’ stratification only in the last three deciles (Figure 3C-Figure 5). The relative importance of the selected 14 features of the Naïve Bayes model is shown on the radar plot in Figure 4.

## Discussion

In this study, we developed and validated a machine learning based risk score (called Piacenza score) to predict the mortality risk among hospitalized patients with COVID-19 pneumonia. This score is based on only six variables readily available variables at hospital admission.

Satisfactory performance, measured in terms of the AUCs in both the testing and external validation cohorts, was achieved with an excellent patient’s stratification.

In crowded hospitals, and with shortages of medical resources, this simple model can help to quickly prioritize patients: if the patient’s estimated risk is low, the clinician may choose to monitor, whereas a high-risk estimate might support aggressive treatment or admission to the ICU. Data from China, Europe and United States reported a hospitalization rates of 20% to 31 %, an ICU admission rates from 17% to 35%, and an in-hospital mortality between 15% and 40% ^12^. In the current study, the in-hospital 30-day mortality was 34% with lower survival for older patients with pre-existing comorbidities and with clinical signs and symptoms suggesting respiratory failure at hospital admission.

In line with previous findings, we found that the most common laboratory abnormalities among patients who died were related to the inflammatory process, renal and liver damage and pro-coagulation status^13-14^

In the presence of a large number of patients requiring intensive care and threating to overwhelm healthcare systems around the world, several models to predict survival and guide clinical decisions in COVID-19 pneumonia were developed. ^15^ However, many of these models have been found to have a high risk of bias, which could reflect the development in small study population with high risk of overfitting and poor generalization properties to unseen cohorts, and without clear details of model derivation and testing^16^.

The recent spread of artificial intelligence brought novel ways to combat current global pandemics by collecting and analysing large amounts of data, identifying trends, stratifying patients on the basis of the risk, and proposing solutions at population level instead of the single individual level. ^17-18^

In the COVID-19 pandemic, machine learning approaches have been used to predict the outbreak, to diagnose the disease, to analyse Chest-X ray and TC-scan images, and more recently to predict mortality or progression risk to severe respiratory failure. ^19-20^

Ye Yuan and colleagues developed a simple prognostic risk score based on a logistic-regression classifier and including three laboratory markers: LDH, high-sensitivity CRP (hs-CRP), and lymphocyte percentage. This score was developed from a cohort of 1’479 patients and externally validated in two independent cohorts reaching an accuracy of 95% in predicting the risk of mortality. However, the model comprised only Chinese patients during the early stages of the outbreak and more importantly it seems to have a significant selection bias as it did not include patients with mild and moderate disease at admission. ^21^

The 4C mortality score, developed and validated by the International Severe Acute Respiratory and Emerging Infections Consortium (ISARIC), based on eight clinical and laboratory variables, achieved an AUC of 0.78 in predicting mortality and it is easy-to-use with a pragmatic design. In fact, to calculate the score, no external tool or complex mathematical equation is required, but results can be immediately inspected bedside. ^22^ However, due to the rapidly evolving of the virus’s characteristics and impact on population, the score should be continuously updated and, for example, the 4C score did not include patients from the second wave of pandemics. At the same, involving a broad range of individuals, it could not be suited for narrower and more specific clinical scenarios, like patients affected by severe pneumonia.

The Piacenza score contains parameters reflecting patient demographics, comorbidity, and physiology at hospital admission. It shares some characteristics with the 4C score such as age, gender, comorbidities, and P/F, but includes also unexplored features like temperature and MCHC deriving from a substantially different variables selection. Unlike traditional scores based on logistic regression analysis mixed with a knowledge-driven approach where a score is assigned by an expert to each of the limited number of selected variables, the proposed predictive model is purely data-driven and it is not affected by a clinically oriented, potentially biased, choice of variables. ^23^

The level of performance of our model is comparable with the 4C mortality score applied to the test cohort used in this paper. However, we remark that 4C mortality score was derived based on a population of 35’000 patients, while the Naïve model providing the Piacenza score was trained using information coming only from 852 patients. This is indicative of the high representativeness of the training cohort considered in our study. Furthermore, although a similar discriminative power between the 4C and the Piacenza score, the latter score showed better performance in stratifying patients according to their mortality risk which is of paramount importance in selecting the appropriate treatment and for resources allocations. We also externally tested our score achieving a good performance and confirming that our data-driven model is robust despite it relies on variables deemed relevant in this context without actually knowing their semantics.

Overall, the Piacenza Score has several advantages: firstly, it relies on objective clinical and laboratory measurements not affected by human interpretation; secondly, it is tested and validated also in patients belonging to the second waves; thirdly, it is automatically generated through a combination of variables widely available at the hospital admission; finally, as opposed to traditional epidemiological predictive models, the Piacenza score has the added advantage of adaptive learning, trend-based recalibration and flexibility. This means that the Piacenza Score could be adapted based on a newer understanding of the disease progression as well as on the impact of the interventions, such as vaccines and newer pharmacological treatments. In fact, Naïve Bayes algorithm, during its learning phase, generates a summary of the dataset where each variable is associated to the outcome in terms of a probabilistic dependence. This summary describes the current dataset’s distribution and can be quickly and easily updated when a new observation is available, adapting itself to the changes inside the population. The Piacenza score is highly flexible as it is confirmed by the result obtained by training the second Naïve Bayes with 14 manually-chosen features, presented in this work. Therefore, if the Piacenza score variables are missing, the physician can still receive a customized result (with respect to the available variables) associated to the best possible accuracy in the specific situation.

Likewise, if new data are available, they can be used to train a new version of the Piacenza Score and study the possible fingerprints of COVID-19 variants.

Finally, the score’s predictors are not chosen a-priori (like, for example, in 4C mortality score), but as the product of a machine learning-based optimization technique, which considers the smallest possible subset of leading predictors associated to the best possible performance

This study has room for further improvement, which is left for future work. Firstly, given that the proposed machine learning method is purely data-driven, our model may vary if a different dataset was used. As more data become available, the model can be refined and performance of the Piacenza score can further increase. To this aim, we are currently looking forward to subsequent large-sample and multi-centred studies. Finally, new variables such as d-dimer and troponin, currently not available, but which are known to be associated with a higher mortality risk in COVID-19 pneumonia may be included in future analysis.

## Data Availability

If request, we agree to publicly share the study data and analysis source code.

## Contributors

GH, DP, MV, MD conceived the study. AB, LR, AB, CM, AN, MM, ES collected the data. MS, UM, AN, DP managed and analysed the data. MM, FP, LR, EM implemented the website. PA and MN provided clinical expertise. MP supervised the work. All authors interpreted the results. GH, MD, MV, DP wrote the manuscript, which was approved by all the authors.

## Data sharing

We agree to publicly share the study data and analysis source code.

## Acknowledgments

None

